# Teaching an ECG model where to look does not make it more robust to missing leads: a pre-registered comparison of guideline-based attention guidance with random lead masking

**DOI:** 10.64898/2026.10.17.26365795

**Authors:** Mohamed Nour Bou Ali, Mohamed Mahmoud

## Abstract

**Background:** Real-world 12-lead electrocardiograms (ECGs) often have missing or corrupted leads. A deep-learning classifier can be prepared for this by random lead masking during training, or by injecting clinical knowledge, for example by pulling its attention towards the leads that guidelines use for each diagnosis. Whether this knowledge adds robustness beyond masking is unknown.

**Methods:** Pre-registered comparison of three arms sharing one network (per-lead 1D-ResNet encoder and per-diagnosis attention over the 12 leads): standard training (M0); random whole-lead masking (M1); masking plus a Kullback-Leibler penalty pulling attention towards a fixed guideline-derived key-lead map (M2, guideline-based attention guidance). Six diagnoses; five seeds per arm. Development on PTB-XL (21,799 ECGs); single sealed evaluation on PTB-XL fold 10 (2,198 ECGs) and on CODE-15% (233,204 exams, external validation). Primary endpoint: AUROC difference M2 − M1, averaged over first-degree AV block, right and left bundle branch block and atrial fibrillation and over 1 to 6 lost (zeroed) leads, with a patient-level paired BCa bootstrap 95% confidence interval.

**Results:** The guidance did not change robustness: ΔAUROC −0.0010 (95% CI −0.0033 to +0.0006), an interval entirely within the pre-specified ±0.02 margin; the result was unchanged with another bootstrap seed, other interval methods and leave-one-diagnosis-out analyses. Masking improved AUROC: M1 − M0 = +0.0134 (95% CI +0.0102 to +0.0172) when one key lead was lost, and the number of lost leads tolerated before AUROC fell by more than 0.02 rose from 7 (M0) to 10 (M1). M2 attention matched the map and M2 was non-inferior on intact ECGs, but its calibration slope was lower for all six diagnoses in both datasets. M2 lost slightly more when all key leads were lost together (−0.0067) and gained only when one to three leads remained (exploratory; AUROC 0.9408 vs 0.9210 with 11 leads lost). CODE-15% reproduced every pattern (primary −0.0007, 95% CI −0.0011 to −0.0003).

**Conclusions:** Random lead masking suffices to make an ECG classifier robust to missing leads; adding guideline knowledge aligns attention with clinical criteria but adds no robustness and worsens calibration. Guideline-aligned attention should not be taken as evidence of robustness.

## 1. Introduction

Deep-learning interpretation of the 12-lead electrocardiogram (ECG) matches cardiologists on good-quality tracings (1,2) and enables new uses (3,4). In practice, however, ECGs are often incomplete: electrodes detach or are misconnected (5), leads are buried in noise, paper tracings are photographed or poorly digitised (6), and devices record fewer leads (7,8). A clinically deployed ECG model must therefore remain reliable when some leads are missing.

Developers have two options. The first is plain data augmentation: random leads are erased during training so that the model learns to do without them (6,7,9). The second injects medical knowledge: for each diagnosis, guidelines name the leads where the criteria are read — V1 and V2 for right bundle branch block, I, aVL, V5 and V6 for left bundle branch block (10). Post-hoc analyses show that networks often rely on these leads spontaneously (11–13), and some architectures already build lead–abnormality knowledge in to improve accuracy (14,15). It is tempting to infer that a model that “looks in the right place” will also be more reliable when leads are lost.

This inference is not self-evident. A plausible explanation is not evidence of reliability (16,17), attention weights are not necessarily the mechanism of a decision (18,19), and attribution methods for ECG models are themselves unstable (20). Above all, to our knowledge, no study has compared a knowledge-guided ECG model with a control receiving **exactly the same masking augmentation**; without such a control, the benefit of masking can be wrongly credited to clinical knowledge.

We therefore tested, in a pre-registered study with sealed evaluation and external validation, whether imposing a guideline-derived key-lead map on a model’s attention makes it more robust to lost leads than random lead masking alone. Hypotheses, fixed before any test-set prediction, were: (H1, primary) the robustness of M2 differs from that of M1 when 1 to 6 leads are lost; (H2) M2 is not inferior to M1 by more than 0.02 AUROC on intact ECGs; (H3) no difference for the two negative-control diagnoses; (H4) the number of lost leads tolerated (k^*^) differs between M2 and M1.

### 2. Methods

### 2.1 Design and pre-registration

Secondary analysis of two public de-identified datasets, reported according to TRIPOD+AI (21) (checklist in the Supplementary Material). The protocol was registered on OSF on 16 October 2026 (doi:10.17605/OSF.IO/M9UR2) before any prediction on PTB-XL fold 10 or CODE-15%; these data could only be read by a locked notebook that required the registration identifier, and they were predicted in a single pass. The 30 training runs and a label-permutation leakage control had been completed beforehand on the development folds only. **Declared deviation:** the original primary endpoint (loss of a single key lead) was replaced, before registration, by the degradation with the number of lost leads, after observing on the validation fold only that losing one lead barely changed AUROC; the original endpoint is reported as a secondary endpoint. Other deviations were technical (the sealed evaluation was split across notebooks; a faster weighted-AUROC algorithm gave bit-identical values) and are listed in the Supplementary Material.

### 2.2 Data

**PTB-XL 1.0.3** (22,23) (Germany; 21,799 10-s ECGs from 18,869 patients) was used for development with its official patient-level folds: folds 1–8 for training (17,418 ECGs), fold 9 for validation (2,183) and fold 10 for the internal test (2,198 ECGs, 1,904 patients). **CODE-15%** (24,25) (Brazil) was used for external validation: the first exam of each patient (233,647), minus 443 7-s tracings, i.e. 233,204 exams. Six diagnoses present in both datasets were studied: first-degree atrioventricular block (AVB1), complete right bundle branch block (RBBB), complete left bundle branch block (LBBB), atrial fibrillation (AF), sinus bradycardia and sinus tachycardia. In PTB-XL a diagnosis was positive when its statement was present, whatever its likelihood (rhythm statements have zero likelihood by construction); in CODE-15% the dataset labels were used. Both datasets also contain normal ECGs and ECGs with other abnormalities (infarction, hypertrophy, repolarisation changes…): all were kept, without selection. Counts are given in Table 1 and the coding of every variable in the Supplementary Material (S0).

**Table 1.** Counts. Number of ECGs and positives per diagnosis. An ECG may carry several diagnoses, and a normal ECG may carry a rhythm statement (sinus bradycardia or tachycardia). ^*^ CODE-15% first exams before exclusion of the 7-s tracings (233,647 exams).

|  | PTB-XL training<br>(folds 1–8) | PTB-XL validation<br>(fold 9) | PTB-XL test (fold<br>10) | CODE-15%<br>(external) |
| --- | --- | --- | --- | --- |
| ECGs (patients) | 17,418 (15,023) | 2,183 (1,942) | 2,198 (1,904) | 233,204 (233,204) |
| Normal ECGs | 7,596 | 955 | 963 | 92,138* |
| At least one of the<br>six diagnoses | 3,506 | 435 | 436 | 22,776* |
| Neither normal<br>nor one of the six<br>(other<br>abnormalities) | 6,858 | 846 | 864 | 118,733* |
| Women | — | — | 1,066 | 138,630 |
| Age ≥ 65 years | — | — | 1,048 | 69,663 |
| AVB1 | 634 | 80 | 79 | 3,366 |
| Complete RBBB | 432 | 55 | 54 | 5,676 |
| Complete LBBB | 428 | 54 | 54 | 3,374 |
| Atrial fibrillation | 1,211 | 151 | 152 | 3,497 |
| Sinus bradycardia | 509 | 64 | 64 | 3,654 |
| Sinus tachycardia | 661 | 83 | 82 | 5,198 |

### 2.3 Pre-processing

PTB-XL signals were resampled from 500 to 400 Hz (their original acquisition rate); CODE-15% signals are recorded at 400 Hz. Each tracing was cut to 10 s (4,000 samples), filtered with a zero-phase 0.5–40 Hz Butterworth band-pass and standardised per lead. The lead order (I, II, III, aVR, aVL, aVF, V1–V6) was verified in both datasets.

### 2.4 Model

The model was developed for this study by the first author, on the principle of residual networks (ResNet) (26); it has 1,040,806 parameters. A shared 1D convolutional encoder (four residual stages with 32, 64, 128 and 128 filters, kernel of 15 samples, 128-dimensional output) processes each lead separately; a learned lead embedding encodes lead identity. For each diagnosis, an attention layer weights the 12 lead representations (positive weights summing to 1) before a sigmoid output. These weights are read directly; no post-hoc attribution method is needed. The network is randomly initialised and trained only on PTB-XL folds 1–8: no pre-trained model is used, so no test ECG (fold 10) and no CODE-15% exam could have been seen before the sealed evaluation.

### 2.5 Training arms and key-lead map

- **M0:** standard training (binary cross-entropy).
- **M1:** M0 + random whole-lead masking: each lead is set to zero with probability 0.25 (at least one lead kept).
- **M2 (guideline-based attention guidance):** M1 + λ × KL(map ‖ attention), computed on positive (ECG, diagnosis) pairs only; this penalty pulls attention towards the key-lead map.

The key-lead map (Table 2, Figure 2A) was derived from guideline texts read in full (10,27–30) and validated by the first author before training (version 1.0). Each lead received a relative weight of 1 (key), 0.3 (secondary) or 0.03 (other) and each row was normalised. For RBBB and LBBB the key leads are those of the guideline criteria; for AVB1 and AF no guideline names a lead, and the choice (II; II and V1) is a clinical convention. The two rate disorders received a uniform map and serve as **negative controls**. Was chosen on fold 9 from {0.1, 0.3, 1, 3} by a pre-specified rule (mean AUROC with one key lead erased): λ^*^ = 0.1. Optimisation used AdamW (31) (learning rate 0.001), batches of 64, at most 40 epochs and early stopping on fold-9 AUROC; seeds 0 to 4 for each arm.

**Table 2.** Guideline-based key-lead map (version 1.0). Relative weights: key 1, secondary 0.3, other 0.03, then row normalisation.

| Diagnosis | Key leads | Secondary leads | Basis |
| --- | --- | --- | --- |
| AVB1 | II | I, aVF, V1 | convention (PR is measured globally (27,28)); sinus P wave (29) |
| Complete RBBB | V1, V2 | I, V5, V6 | guideline criteria (10) |
| Complete LBBB | I, aVL, V5, V6 | V1, V2, V3 | guideline criteria (10) |
| Atrial fibrillation | II, V1 | I, aVF | convention (no lead named (30)); P wave (29) |
| Sinus bradycardia and tachycardia | uniform | — | negative controls |

**Figure 1.**
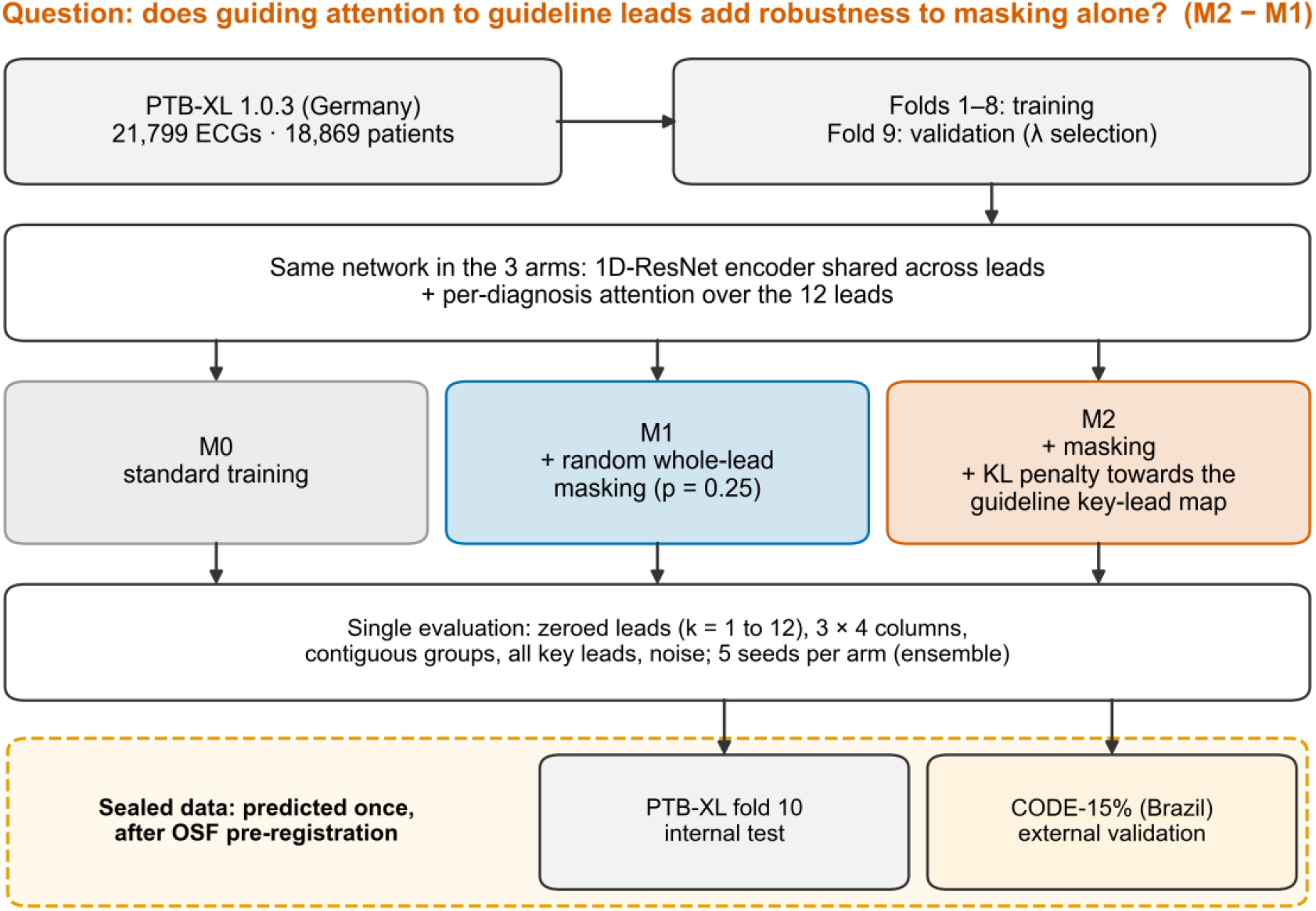
Study design. The three arms share the same network; only training differs. PTB-XL fold 10 and CODE-15% were predicted once, after pre-registration.

**Figure 2.**
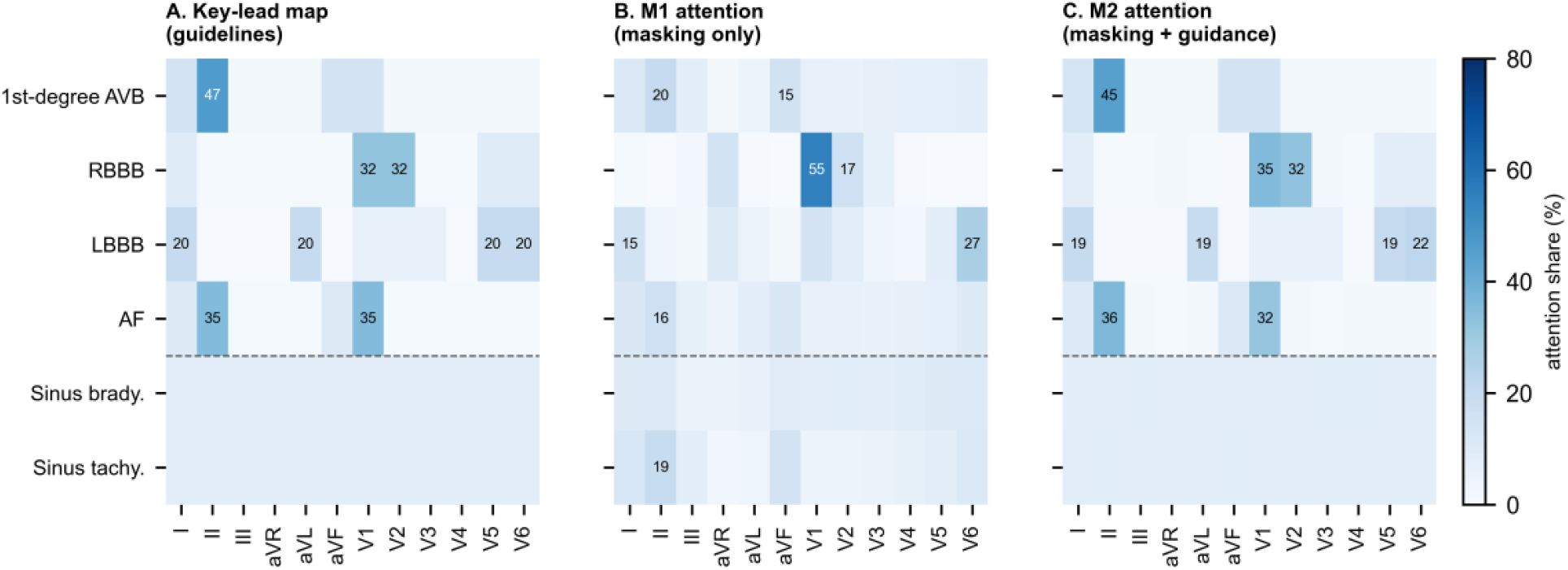
Key-lead map and learned attention (PTB-XL fold 10, positive ECGs). A, guideline-based key-lead map; B, M1 attention; C, M2 attention (%). Below the dashed line: negative controls (uniform map).

### 2.6 Test scenarios

Applied identically and pairwise to all arms in a single pass: intact ECG; loss (zeroing) of k leads for k = 1 to 12 (all combinations for k = 1 and 11; 10 subsets drawn once for k = 2 to 10); the four 3 × 4 paper-layout columns; the contiguous lead groups of the Fourth Universal Definition of Myocardial Infarction (32,33); loss of all key leads of a diagnosis; one key or accessory lead; white noise on one lead (PTB-XL only).

### 2.7 Endpoints and statistical analysis

Each arm’s prediction is the mean probability of its five seeds (34). For each diagnosis, AUROC compares the positive ECGs with all other ECGs of the dataset (normal ECGs and other abnormalities). **Primary endpoint:** Δ = mean over AVB1, RBBB, LBBB and AF and over k = 1 to 6 of AUROC_M2(k) − AUROC_M1(k) on PTB-XL fold 10, with a 95% patient-level paired bootstrap confidence interval (2,000 resamples, bias-corrected and accelerated (BCa) (35), seed 42); H1 is supported if the CI excludes 0. **Pre-specified secondary endpoints:** Δ(k) for each k (Holm correction (36)); k^*^, the smallest k at which AUROC falls by more than 0.02; paper columns, contiguous groups and all key leads; one key lead (original endpoint; paired DeLong test (37) per diagnosis); non-inferiority on intact ECGs (margin 0.02); calibration (slope, expected calibration error, Brier score (38,39); a slope below 1 indicates probabilities too close to 0 or 1); robustness to noise; attention concordance with the map; descriptive sex and age subgroups; external validation (CODE-15%, jackknife over 200 patient blocks); the likelihood ≥ 50 labelling rule. **Post hoc analyses** (not pre-registered and labelled as such): equivalence reading by two one-sided tests (40) with the pre-specified margin, other interval methods and bootstrap seed, per-diagnosis decomposition, seed-by-seed analyses and an independent re-computation of all endpoints.

## 3. Results

### 3.1 Population and checks

Counts are shown in Table 1. Fold 10 contained 963 normal ECGs (2,198 in total), 436 ECGs with at least one of the six diagnoses and 864 ECGs with other abnormalities (a normal ECG may also carry a rhythm statement); among CODE-15% first exams, 92,138 of 233,647 were normal. On fold 10, positives for the four primary diagnoses ranged from 54 to 152; in CODE-15%, from 3,366 to 5,676. The leakage control was negative: with permuted labels, validation AUROC stayed at chance level (mean 0.534). The independent re-computation reproduced the primary endpoint and its bounds exactly; 38 of 38 integrity checks passed. No bootstrap resample had to be redrawn.

### 3.2 Primary endpoint

On fold 10, Δ = **−0.0010** (95% BCa CI −0.0033 to +0.0006; p = 0.291); the CI included 0, so H1 was not supported (Figure 3C, Table 3). *Post hoc*, the 90% CI (−0.0028 to +0.0004) lay entirely within the ±0.02 margin, which meets the equivalence criterion. The result did not change with another bootstrap seed (CI −0.0032 to +0.0007), with percentile (−0.0031 to +0.0008) or normal (−0.0030 to +0.0009) intervals, or when any one diagnosis was removed (all intervals included 0; Supplementary Material). The negative sign came mostly from LBBB (Δ −0.0033); without LBBB, Δ = −0.0003. The likelihood ≥ 50 rule changed no fold-10 positive and therefore gave an identical result.

**Table 3.** AUROC differences M2 − M1 [95% BCa CI] (five-seed ensemble).

| Scenario | PTB-XL fold 10 | CODE-15% |
| --- | --- | --- |
| **Primary: 1 to 6 lost leads** | **–0.0010 [–0.0033, +0.0006]** | **–0.0007 [–0.0011, –0.0003]** |
| One key lead lost | +0.0008 [–0.0007, +0.0023] | +0.0001 [–0.0002, +0.0005] |
| One accessory lead lost | +0.0009 [–0.0006, +0.0025] | +0.0012 [+0.0008, +0.0015] |
| 3 × 4 paper columns | –0.0020 [–0.0044, –0.0001] | –0.0017 [–0.0021, –0.0013] |
| Contiguous lead groups | –0.0019 [–0.0044, –0.0001] | –0.0029 [–0.0034, –0.0025] |
| All key leads lost | –0.0067 [–0.0100, –0.0039] | –0.0063 [–0.0069, –0.0057] |
| Intact ECG, six diagnoses (non-inferiority) | –0.0012 [–0.0039, +0.0004] | +0.0007 [+0.0005, +0.0010] |
| *Effect of masking, M1 – M0 (one key lead)* | +0.0134 [+0.0102, +0.0172] | +0.0152 [+0.0149, +0.0156] |
| k* M1 / M2 (lost leads) | 10 / 11 | 9 / 11 |

**Figure 3.**
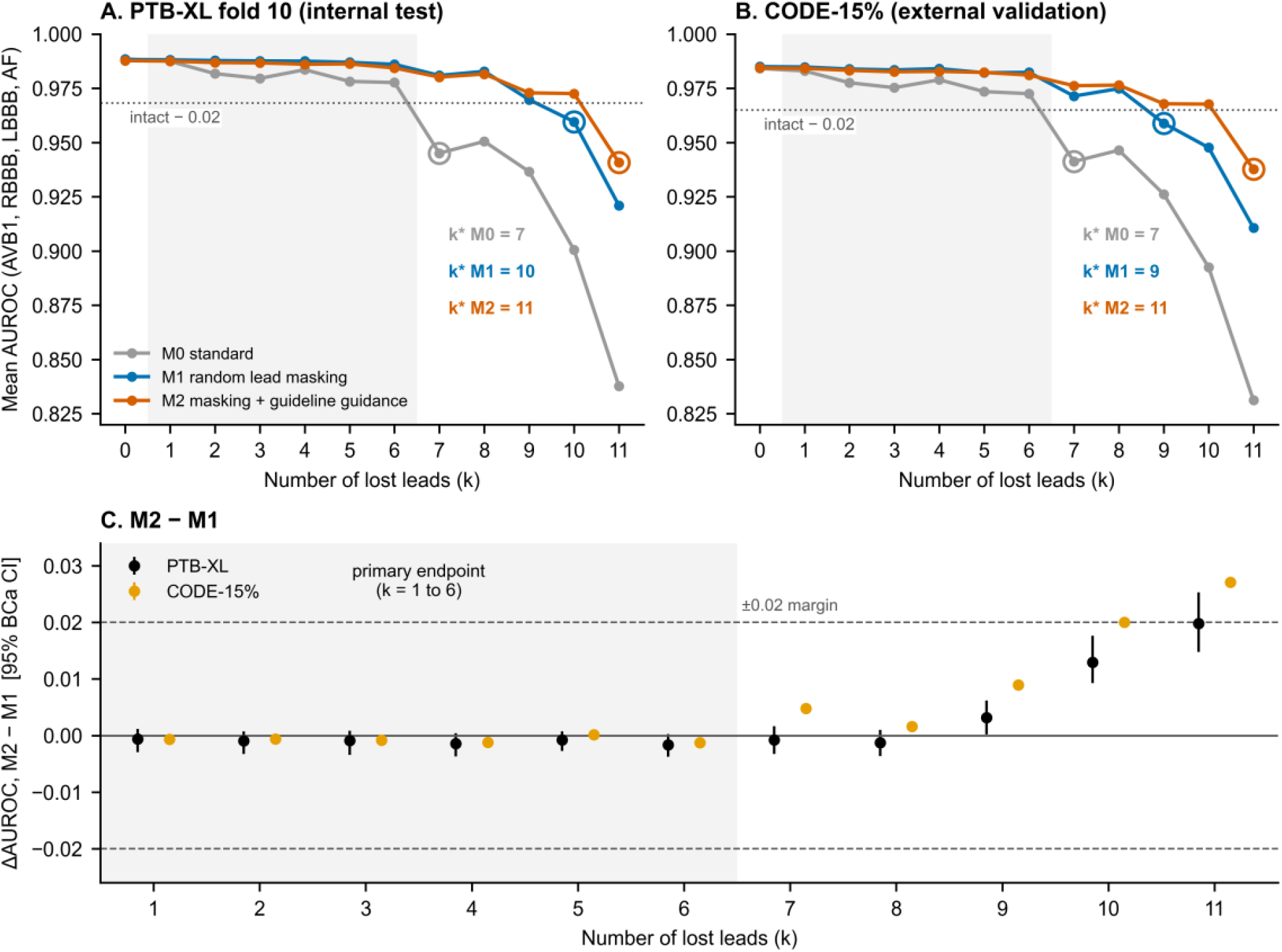
Degradation with the number of lost leads. A–B, mean AUROC of the four primary diagnoses; dotted line: intact AUROC − 0.02; rings: k^*^, the first k below this threshold; grey band: range of the primary endpoint. C, Δ(k) = M2 − M1 with 95% BCa CI (CODE-15% intervals are smaller than the markers).

### 3.3 Effect of masking

When one key lead was lost, masking improved AUROC by +0.0134 (95% CI +0.0102 to +0.0172), whereas the guidance added only +0.0008 (−0.0007 to +0.0023), about 6% of the masking gain (Figure 4A). Without masking, AUROC fell by more than 0.02 from k^*^ = 7 lost leads, against 10 for M1 and 11 for M2 (Figure 3A). With 6 leads lost, mean AUROC was 0.9777 (M0), 0.9860 (M1) and 0.9844 (M2), compared with 0.9883 on intact ECGs (M1).

**Figure 4.**
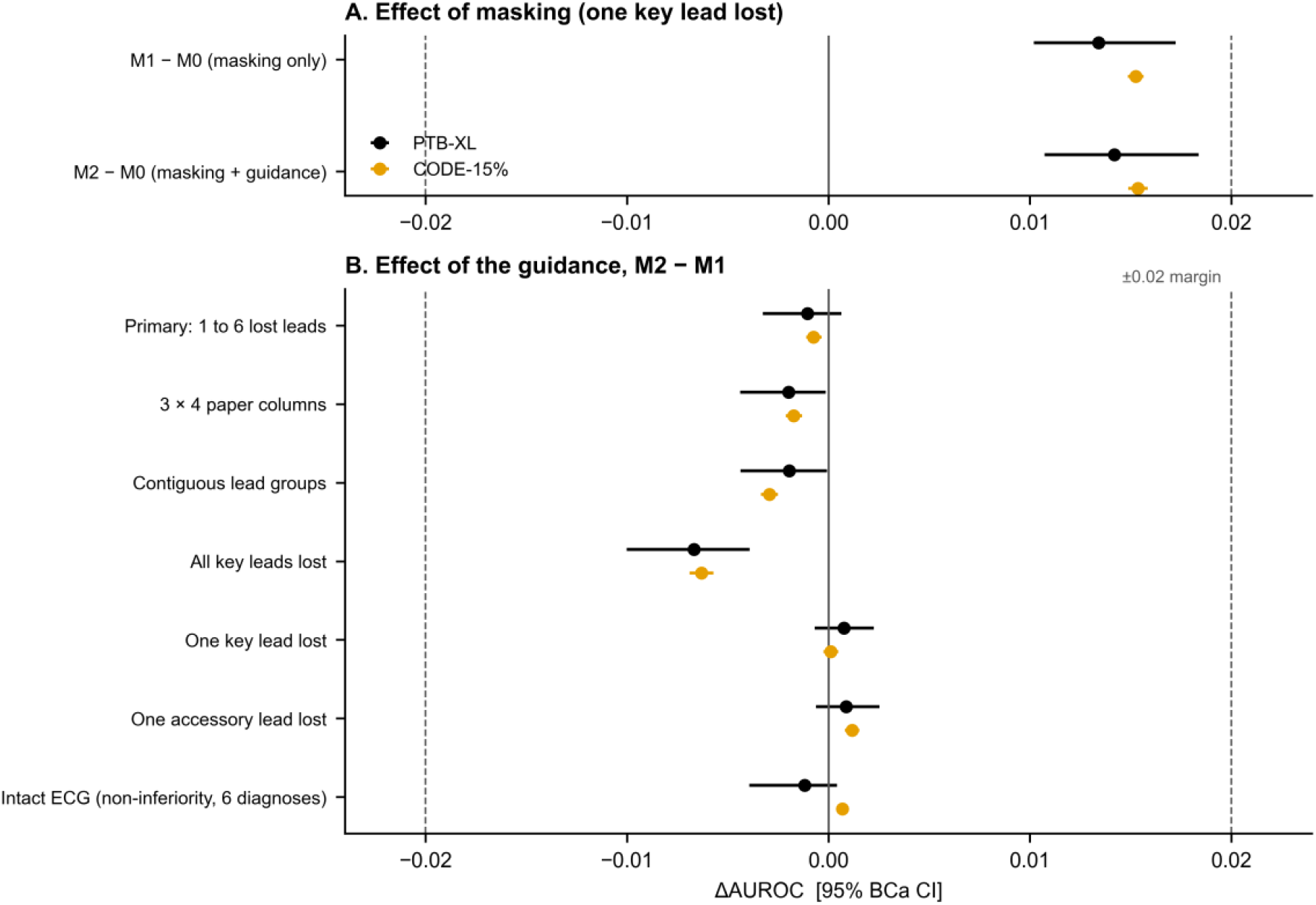
Effect of masking and effect of the guidance. A, masking gain when one key lead is lost. B, all M2 − M1 differences; dashed lines: ±0.02 margin.

### 3.4 Grouped lead losses

When leads were lost in blocks, Δ was negative (Figure 4B): paper columns, Δ = −0.0020; contiguous groups, −0.0019; V1–V6 group, −0.0035; **all key leads lost together, −0.0067** (95% CI −0.0100 to −0.0039). None of these differences reached 0.007 AUROC.

### 3.5 Extreme losses (exploratory analysis)

For k = 1 to 8, Δ(k) ranged from −0.0016 to −0.0006; no value was significant after Holm correction. Δ became positive when only one to three leads remained: Δ(9) = +0.0032 (nominal p = 0.032, Holm-adjusted 0.32), Δ(10) = +0.0130 and Δ(11) = +0.0198 (Holm-adjusted bootstrap p < 0.001); with 11 leads lost, mean AUROC rose from 0.9210 (M1) to 0.9408 (M2). Accordingly, k^*^ was one lead higher (percentile interval 1 to 2).

### 3.6 Attention, intact-ECG discrimination and calibration

The guidance changed attention (Figure 2): the share of M2 attention on key leads approached that of the map (AF: 0.679 vs 0.704 for the map and 0.220 for M1; AVB1: 0.450 vs 0.198), and the divergence between attention and map fell more than 100-fold (M2/M1 ratio 0.0042 to 0.0093). For RBBB, M1 already attended to V1–V2 more than the map did (0.719 vs 0.643).

Negative controls (uniform map), one lead lost: in PTB-XL, Δ = −0.0047 for sinus bradycardia (95% CI −0.0110 to −0.0013; adjusted DeLong p = 0.51) and −0.0057 for sinus tachycardia (−0.0260 to +0.0001); in CODE-15%, +0.0001 (−0.0002 to +0.0004) and +0.00003 (−0.00026 to +0.00033).

On intact ECGs, M2 was non-inferior to M1 (Δ over six diagnoses −0.0012, 95% CI −0.0039 to +0.0004). However, **the calibration slope of M2 was lower than that of M1 for all six diagnoses** (difference −0.3513 to −0.1310; Figure 5; AF: slope 0.533 vs 0.664). Brier scores changed little (difference −0.0018 to +0.0017).

**Figure 5.**
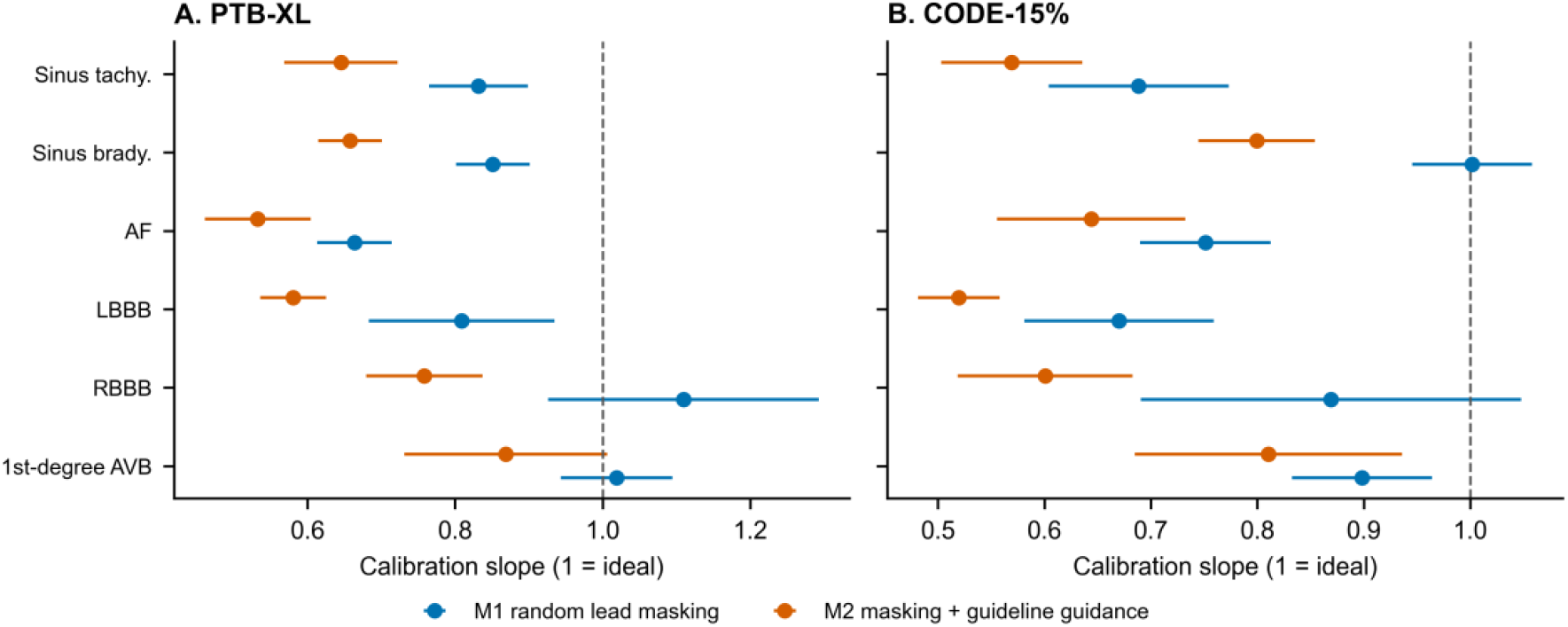
Calibration on intact ECGs. Calibration slope (mean ± SD over five seeds); 1 = ideal calibration; a slope below 1 means over-extreme probabilities.

### 3.7 External validation in CODE-15%

In the 233,204 Brazilian exams, the primary endpoint was Δ = −0.0007 (95% CI −0.0011 to −0.0003), an interval entirely within ±0.02. Masking gain: +0.0152; guidance gain: +0.0001. k^*^: 7 (M0), 9 (M1), 11 (M2). All key leads lost: −0.0063. With 11 leads lost: 0.9107 (M1) vs 0.9377 (M2). The calibration slope of M2 was lower for all six diagnoses (difference −0.2686 to −0.0878). The Δ(k) curves of the two datasets were correlated (*post hoc* Spearman correlation 0.78); per-diagnosis single-key-lead differences had the same sign in both datasets for 1 of 6 diagnoses.

## 4. Discussion

### 4.1 Main finding

Teaching an ECG model where to look, according to the guidelines, does not make it more robust to missing leads than random lead masking alone. This null result is **informative**: it was pre-registered, obtained on sealed data, reproduced on an external dataset from another continent and estimated precisely enough to exclude any clinically useful difference (41,42). Robustness comes from masking, which raised k^*^ from 7 to 10 lost leads in PTB-XL and from 7 to 9 in CODE-15% and explains almost all the gain. Without the M1 control, the M2 − M0 gain (+0.0142) would have been credited to clinical knowledge. In CODE-15%, the primary interval excludes 0 only because of the sample size: an effect of a few ten-thousandths of AUROC is clinically negligible, so external validation confirms the PTB-XL result. Finally, testing used unselected ECGs, normal or with other abnormalities, not only abnormal tracings.

### 4.2 Why the knowledge does not help

Our explanation, which was not tested directly, is ECG redundancy: the six limb leads carry only two independent signals and neighbouring precordial leads are correlated. Masking teaches the model to recover the missing information from the remaining leads. The guidance, by contrast, concentrates attention on a few leads: this costs nothing when only some of them disappear but becomes a handicap when all disappear together — exactly what we observed (Δ −0.0067 in PTB-XL and −0.0063 in CODE-15%). Conversely, when only one to three leads remain, knowledge seems to act as a guide where the data no longer suffice; this signal, reproduced in both datasets but outside the primary endpoint, remains a hypothesis. Such a mismatch between a model’s apparent reasoning and its behaviour under distribution shift is a known pitfall of deep learning (43).

RBBB is instructive: without any constraint, M1 already placed more attention on V1–V2 than the map did. The network had found the guideline criterion on its own, in line with post-hoc analyses (11,12). Changing *where* the model looks did not change *what protects it*: attention is not the mechanism of robustness, consistent with the debate on the explanatory value of attention (18,19).

### 4.3 Relation to previous work

Studies that build lead–diagnosis knowledge into ECG networks aimed at accuracy (14,15); studies that use lead masking do not inject per-diagnosis knowledge (6,9). Our study links the two with a control that isolates the effect of knowledge. It extends the “right for the right reasons” principle (44) by measuring what an explanation constraint brings (nothing for robustness) and what it costs (calibration). Lead-selection studies (7,45) address fixed reduced-lead sets; we address the unexpected loss of any lead.

### 4.4 Implications

**For clinicians and for those who evaluate or procure ECG AI tools:** an attention map that “looks at the right leads” does not guarantee reliability on an incomplete tracing; a curve of performance against the number of lost leads, with k^*^, should be requested. M2 probabilities were too close to 0 or 1 (calibration slope below 1); if a tool displays probabilities, local recalibration is required, especially when an explanation constraint was used in training (38). **For developers:** random lead masking is simple, free and sufficient; any gain credited to clinical knowledge should be compared with a control receiving the same augmentation. The scenarios and paired statistics used here form a test bench that transfers to any ECG model.

### 4.5 Strengths and limitations

Strengths: pre-registration, single sealed evaluation, masked control, five seeds, large external validation, leakage control, independent re-computation. Limitations: (1) the primary endpoint was changed before registration after inspection of the validation fold only; (2) AUROCs near 1 leave little room for gains; (3) simulated loss (zeroing) does not represent every real loss (lead reversal, intermittent noise, digitisation artefacts); (4) the map, validated by the first author alone before training, rests on convention for AVB1 and AF, and another map or λ might behave differently; (5) fold-10 positives are few, so per-diagnosis and subgroup analyses are imprecise; (6) CODE-15% labels are partly automatic; (7) calibration differences have no confidence intervals; (8) a single architecture was studied; (9) the uniform map of the negative controls also constrains attention, so they are not perfectly neutral controls, which may explain the small sinus-bradycardia difference in PTB-XL, not found in CODE-15%.

### 4.6 Future work

Test the guidance on genuine reduced-lead recordings (one to three leads), where the exploratory signal appeared; compare other forms of knowledge (measured criteria rather than leads); evaluate “masking plus recalibration” in real-world conditions.

## 5. Conclusions

Random lead masking during training is sufficient to make an ECG classifier robust to missing leads. Adding guideline knowledge makes attention more consistent with clinical criteria, without any gain in robustness and at the cost of worse calibration. Guideline-consistent attention should not be taken as evidence of robustness.

## Supporting information

TRIPOD-AI_checklist

## Data Availability

All data analysed in this study are publicly available. PTB-XL v1.0.3 is available at https://physionet.org/content/ptb-xl/1.0.3/ and CODE-15% at https://zenodo.org/records/4916206. The study protocol was pre-registered on OSF (doi:10.17605/OSF.IO/M9UR2).

https://zenodo.org/records/4916206

https://doi.org/10.17605/OSF.IO/M9UR2

## Declarations

### Ethics

Secondary analysis of public de-identified data (CC BY 4.0); publication of PTB-XL was approved by the PTB ethics committee (PTB-2020-1). No participant was recruited and no additional ethical approval was required.

### Data and code availability

PTB-XL (PhysioNet, doi:10.13026/kfzx-aw45) (46) and CODE-15% (Zenodo, doi:10.5281/zenodo.4916206) are public. The code (MIT licence), configuration and every reported value will be released with the article and are available from the corresponding author on reasonable request. Protocol: doi:10.17605/OSF.IO/M9UR2.

### CRediT

M.N.B.A.: conceptualisation, methodology, software, formal analysis, writing – original draft. M.M.: writing – review & editing (clinical cardiology review).

### Declaration of generative AI and AI-assisted technologies in the manuscript preparation process

During the preparation of this work the authors used Claude (Anthropic) to help refine and test the analysis code and to assist with drafting and language editing of the manuscript. After using this tool, the authors reviewed and edited the content as needed and take full responsibility for the content of the published article.

## Supplementary Material (separate file)

coding of variables, deviations from protocol, lead-loss scenarios in detail, λ selection, interval methods and bootstrap seeds, seed-by-seed analysis, leave-one-diagnosis-out, per-diagnosis results, noise robustness, subgroups, TRIPOD+AI checklist.

