## Supplementary_Material for "Teaching an ECG model where to look does not make it more robust to missing leads: a pre-registered comparison of guideline-based attention guidance with random lead masking"

All values are injected from the result files of the sealed evaluation (`results/`); analyses marked *post hoc* were not pre-registered (OSF doi:10.17605/OSF.IO/M9UR2).

### S0. Coding of variables

| Variable | Dataset | Original coding | Coding in this study |
|---|---|---|---|
| Patient | PTB-XL, CODE-15% | `patient_id` | bootstrap resampling unit |
| Fold | PTB-XL | `strat_fold` 1 to 10 (each patient in one fold) | 1–8 training, 9 validation, 10 test |
| Exam kept | CODE-15% | `timey` filled for the patient's first exam | one exam per patient |
| Tracing length | CODE-15% | 10 s or 7 s (zero-padded) | 7-s tracings excluded |
| AVB1, RBBB, LBBB, AF, sinus bradycardia and tachycardia | PTB-XL | `scp_codes`: 1AVB, CRBBB, CLBBB, AFIB, SBRAD, STACH with a likelihood of 0–100 | 1 if the statement is present, whatever its likelihood, else 0; sensitivity: likelihood ≥ 50 for 1AVB, CRBBB, CLBBB |
| Same diagnoses | CODE-15% | Boolean columns `1dAVb`, RBBB, LBBB, AF, SB, ST | 1 / 0 |
| Normal ECG | PTB-XL; CODE-15% | NORM statement; `normal_ecg` column | description only (not a model target) |
| Sex | PTB-XL; CODE-15% | `sex` (0 male, 1 female); `is_male` (Boolean) | female / male (subgroups) |
| Age | PTB-XL; CODE-15% | years (PTB-XL ages > 89 coded 300) | < 65 / ≥ 65 years (code 300 counted ≥ 65) |
| Signal | PTB-XL; CODE-15% | 12 leads (I, II, III, aVR, aVL, aVF, V1–V6), 500 Hz; 400 Hz | 400 Hz, 4,000 samples, filtered, standardised per lead |
| Lost lead | — | — | lead signal set to zero |

| Variable | Dataset | Original coding | Coding in this study |
|---|---|---|---|
| Model output | — | — | probability 0–1 per diagnosis; mean of five seeds |
| Attention | — | — | 12 positive weights summing to 1, per diagnosis and ECG |

### S1. Deviations from the registered protocol

| Date | Deviation | Timing relative to test-set prediction | Effect |
|---|---|---|---|
| 16 Oct 2026 | Primary endpoint changed from “loss of one key lead” to “mean over 1–6 lost leads”, after seeing on fold 9 (validation) that one lost lead barely changed AUROC | before registration and before any test prediction | original endpoint reported as secondary endpoint 4 |
| 16 Oct 2026 | Contiguous-lead-group scenarios added (secondary) | before registration | none on the primary endpoint |
| 16 Oct 2026 | Sealed evaluation re-launched after a technical failure that produced no result; CODE-15% evaluation split into notebooks of six archives | during the sealed pass | none (no prediction had been read) |
| 16 Oct 2026 | Faster exact weighted-AUROC algorithm in the statistics code | after prediction, before statistics | values bit-identical to the registered algorithm (unit test with strict equality) |
| 17 Oct 2026 | Technical note: the registered BCa code adds $10^{-12}$ to the denominator of the acceleration | — | bounds shifted by at most 0.00000005 AUROC; invisible at four decimals |

### S2. λ selection on fold 9 (pre-specified rule)

Mean fold-9 AUROC of the four primary diagnoses with each key lead erased in turn, averaged over five seeds.

| λ | Mean AUROC | SD between seeds |
|---|---|---|
| 0.1 (selected) | 0.9845 | 0.0013 |
| 0.3 | 0.9833 | 0.0010 |
| 1 | 0.9826 | 0.0018 |
| 3 | 0.9827 | 0.0007 |

### S3. Primary endpoint: interval methods, bootstrap seed, labelling rule (*post hoc* except the registered BCa 95%)

| Analysis | Estimate | 95% interval | 90% BCa interval |
|---|---|---|---|
| Registered: BCa, seed 42 | −0.0010 | −0.0033 to +0.0006 | −0.0028 to +0.0004 |
| Independent re-computation, BCa | −0.0010 | −0.0033 to +0.0006 | — |
| Percentile | — | −0.0031 to +0.0008 | — |
| Normal | — | −0.0030 to +0.0009 | — |
| Other bootstrap seed, BCa | −0.0010 | −0.0032 to +0.0007 | −0.0028 to +0.0004 |
| Likelihood ≥ 50 rule (secondary 12) | −0.0010 | −0.0033 to +0.0006 | — |

The likelihood ≥ 50 rule changed no fold-10 positive (AVB1 79, RBBB 54, LBBB 54 under both rules): the sensitivity analysis is identical to the primary analysis by construction and therefore uninformative.

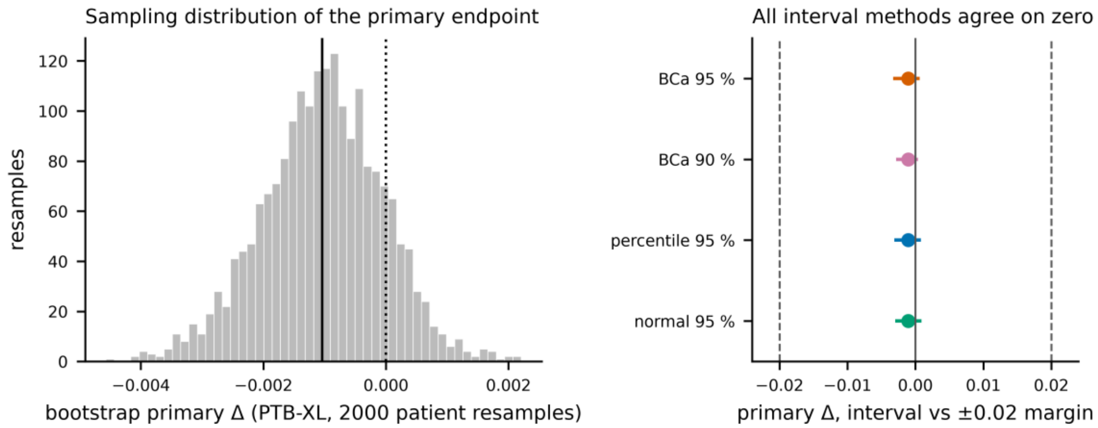


*Bootstrap distribution of the primary endpoint and interval methods.*

### S4. Primary endpoint by diagnosis and leave-one-diagnosis-out (*post hoc*)

| Diagnosis | Δ (mean of k = 1–6) [95% BCa CI] | Primary endpoint without this diagnosis [95% BCa CI] |
|---|---|---|
| AVB1 | +0.0002 [−0.0039, +0.0047] | −0.0014 [−0.0041, +0.0003] |
| RBBB | −0.0007 [−0.0019, +0.0005] | −0.0012 [−0.0041, +0.0011] |
| LBBB | −0.0033 [−0.0127, −0.0009] | −0.0003 [−0.0025, +0.0020] |
| AF | −0.0003 [−0.0048, +0.0046] | −0.0013 [−0.0039, +0.0003] |

Four unplanned analyses without multiplicity correction; the LBBB interval is asymmetric (54 positives).

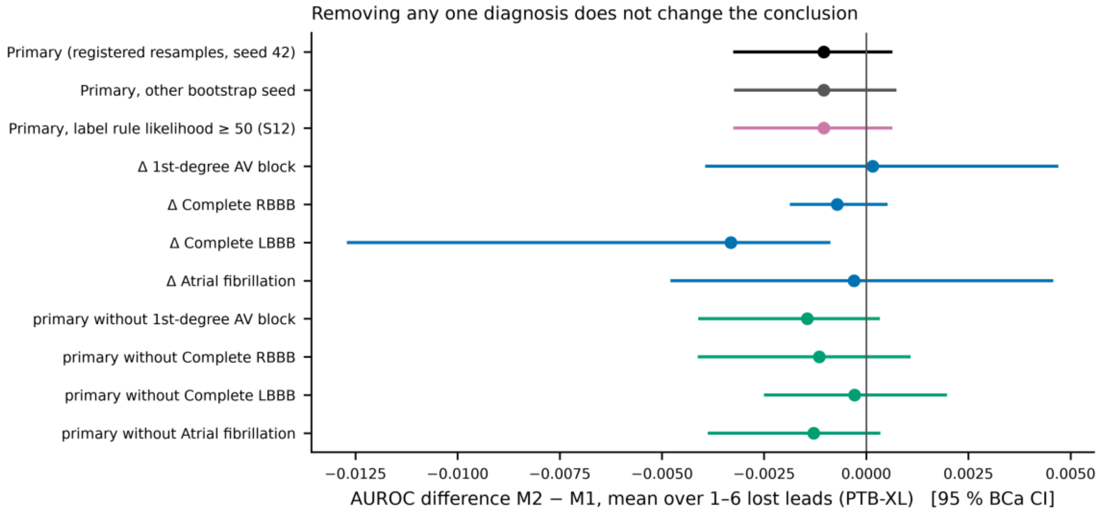


*Decomposition of the primary endpoint.*

### S5. Seed-by-seed analysis (*post hoc*)

The primary endpoint uses the five-seed ensemble, as registered. Comparing M1 seed s with M2 seed s gives Δ from −0.0006 to +0.0019; over the 25 seed pairs Δ ranges from −0.0010 to +0.0027 (7 negative). All values are far below 0.02. A plausible but untested explanation for the slightly positive single-seed and slightly negative ensemble values is that the guidance makes the five M2 seeds more similar, so that averaging them helps M2 less than M1.

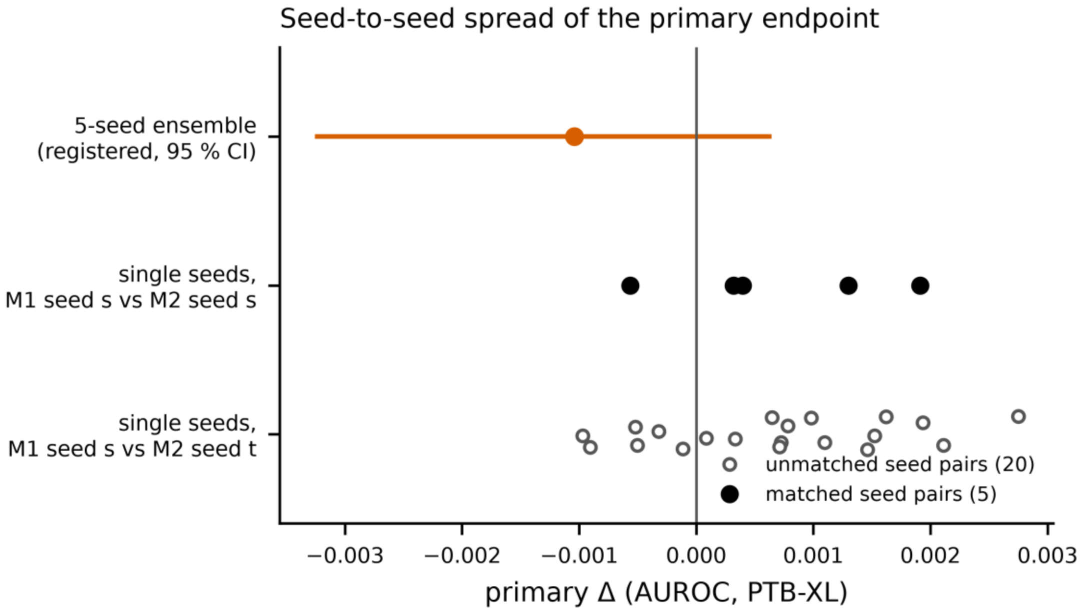


*Seed-by-seed primary endpoint.*

### S6. Δ(k) for each number of lost leads (secondary endpoint 1)

| k | PTB-XL Δ [95% BCa CI] | Holm p | CODE-15% Δ [95% BCa CI] | Holm p |
|---|---|---|---|---|
| 1 | −0.0006 [−0.0029, +0.0012] | 1.000 | −0.0007 [−0.0011, −0.0002] | 0.004 |
| 2 | −0.0010 [−0.0033, +0.0007] | 1.000 | −0.0006 [−0.0010, −0.0002] | 0.004 |
| 3 | −0.0009 [−0.0034, +0.0008] | 1.000 | −0.0008 [−0.0012, −0.0004] | < 0.001 |
| 4 | −0.0014 [−0.0037, +0.0004] | 1.000 | −0.0012 [−0.0016, −0.0008] | < 0.001 |
| 5 | −0.0008 [−0.0027, +0.0008] | 1.000 | +0.0001 [−0.0002, +0.0005] | 0.477 |

| k | PTB-XL Δ [95% BCa CI] | Holm p | CODE-15% Δ [95% BCa CI] | Holm p |
|---|---|---|---|---|
| 6 | −0.0016 [−0.0038, +0.0002] | 0.828 | −0.0012 [−0.0016, −0.0008] | < 0.001 |
| 7 | −0.0008 [−0.0033, +0.0017] | 1.000 | +0.0048 [+0.0043, +0.0053] | < 0.001 |
| 8 | −0.0012 [−0.0036, +0.0010] | 1.000 | +0.0016 [+0.0012, +0.0021] | < 0.001 |
| 9 | +0.0032 [+0.0002, +0.0062] | 0.320 | +0.0090 [+0.0084, +0.0095] | < 0.001 |
| 10 | +0.0130 [+0.0093, +0.0176] | < 0.001 | +0.0200 [+0.0194, +0.0207] | < 0.001 |
| 11 | +0.0198 [+0.0148, +0.0253] | < 0.001 | +0.0271 [+0.0264, +0.0278] | < 0.001 |
| 12 | +0.0011 [−0.0014, +0.0112] | 1.000 | 0.0000 [−0.0001, 0.0000] | 0.142 |

k = 12 (all leads zeroed) is an internal control: AUROC is close to 0.5 in all arms. Holm p-values of 0 mean that no bootstrap resample fell on the other side of zero (reported as < 0.001).

### S7. One key lead lost, by diagnosis (secondary endpoint 4)

AUROC averaged over seeds and over each key lead erased in turn; Δ = M2 − M1 with 95% BCa CI; DeLong test on the seed- and scenario-averaged scores, Holm-adjusted over six diagnoses. SB and ST are negative controls.

| Diagnosis | PTB-XL positives | PTB-XL Δ [95% CI] | Holm p | CODE-15% positives | CODE-15% Δ [95% CI] | Holm p |
|---|---|---|---|---|---|---|
| AVB1 | 79 | +0.0026 [−0.0017, +0.0074] | 1.000 | 3,366 | −0.0011 [−0.0018, −0.0005] | < 0.001 |
| RBBB | 54 | +0.0001 [−0.0011, +0.0014] | 1.000 | 5,676 | −0.0036 [−0.0045, −0.0028] | < 0.001 |
| LBBB | 54 | −0.0007 [−0.0027, +0.0005] | 0.822 | 3,374 | +0.0028 [+0.0022, +0.0034] | < 0.001 |
| AF | 152 | +0.0010 | 1.000 | 3,497 | +0.0024 | < 0.001 |

| Diagnosis | PTB-XL positives | PTB-XL Δ [95% CI] | Holm p | CODE-15% positives | CODE-15% Δ [95% CI] | Holm p |
|---|---|---|---|---|---|---|
| | | [−0.0030, +0.0049] | | | [+0.0018, +0.0032] | |
| SB | 64 | −0.0047 [−0.0110, −0.0013] | 0.511 | 3,654 | +0.0001 [−0.0002, +0.0004] | 0.996 |
| ST | 82 | −0.0057 [−0.0260, +0.0001] | 1.000 | 5,198 | 0.0000 [−0.0003, +0.0003] | 0.996 |

In CODE-15% some per-diagnosis differences have small p-values because of the sample size, but their sign agreed with PTB-XL for only 1 of 6 diagnoses (*post hoc*): they are not reproducible and are not interpreted.

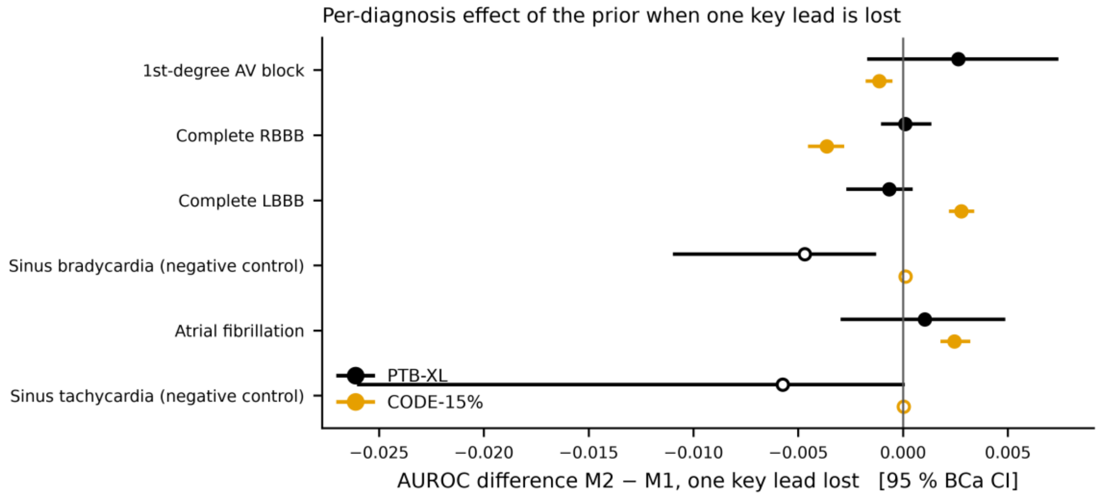


*Per-diagnosis effect of the guidance when one key lead is lost.*

### S8. Calibration on intact ECGs (secondary endpoint 7; mean over five seeds)

| Diagnosis | PTB-XL slope M1 | PTB-XL slope M2 | PTB-XL ECE M1 / M2 | CODE-15% slope M1 | CODE-15% slope M2 | CODE-15% ECE M1 / M2 |
|---|---|---|---|---|---|---|
| AVB1 | 1.019 | 0.869 | 0.0149 / 0.0120 | 0.898 | 0.810 | 0.0061 / 0.0056 |
| RBBB | 1.110 | 0.758 | 0.0054 / 0.0049 | 0.869 | 0.601 | 0.0085 / 0.0081 |
| LBBB | 0.809 | 0.581 | 0.0061 / | 0.670 | 0.519 | 0.0047 / |

| Diagnosis | PTB-XL slope M1 | PTB-XL slope M2 | PTB-XL ECE M1 / M2 | CODE-15% slope M1 | CODE-15% slope M2 | CODE-15% ECE M1 / M2 |
|---|---|---|---|---|---|---|
| | | | 0.0068 | | | 0.0057 |
| AF | 0.664 | 0.533 | 0.0092 / 0.0091 | 0.751 | 0.644 | 0.0148 / 0.0089 |
| SB | 0.851 | 0.658 | 0.0090 / 0.0128 | 1.002 | 0.799 | 0.0102 / 0.0182 |
| ST | 0.832 | 0.645 | 0.0050 / 0.0066 | 0.689 | 0.569 | 0.0080 / 0.0085 |

No confidence intervals were pre-specified for these differences: the finding is descriptive, but consistent in 12 of 12 diagnosis × dataset comparisons.

### S9. Attention concordance with the key-lead map (secondary endpoint 9)

Share of attention on key leads among positive ECGs.

| Diagnosis | Map | PTB-XL M1 | PTB-XL M2 | CODE-15% M1 | CODE-15% M2 | KL ratio M2/M1 (PTB-XL / CODE-15%) |
|---|---|---|---|---|---|---|
| AVB1 | 0.467 | 0.198 | 0.450 | 0.213 | 0.453 | 0.0042 / 0.0048 |
| RBBB | 0.643 | 0.719 | 0.678 | 0.678 | 0.667 | 0.0093 / 0.0093 |
| LBBB | 0.792 | 0.545 | 0.796 | 0.530 | 0.789 | 0.0062 / 0.0077 |
| AF | 0.704 | 0.220 | 0.679 | 0.227 | 0.687 | 0.0058 / 0.0039 |

### S10. Robustness to noise (secondary endpoint 8, PTB-XL)

Normalised area under the AUROC-versus-SNR curve (noise on one key lead), M2 − M1: AVB1 +0.0026, RBBB +0.0018, LBBB −0.0010, AF +0.0018, sinus bradycardia −0.0044, sinus tachycardia −0.0060. No consistent pattern.

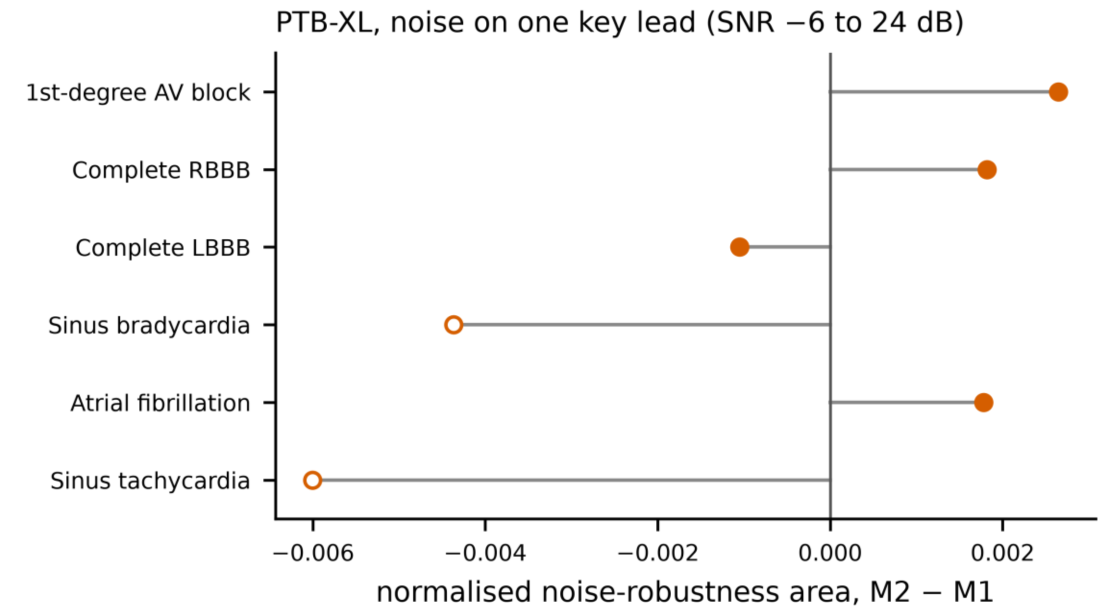


*Robustness to noise.*

### S11. Subgroups (secondary endpoint 10, descriptive, no test)

Mean over the four primary diagnoses of M2 − M1 over 1–6 lost leads.

| Dataset | Women | Men | < 65 years | ≥ 65 years |
|---|---|---|---|---|
| PTB-XL | −0.0020 (n = 1,066) | −0.0002 (n = 1,132) | −0.0010 (n = 1,150) | −0.0017 (n = 1,048) |
| CODE-15% | −0.0002 (n = 138,630) | −0.0015 (n = 94,574) | −0.0007 (n = 163,541) | −0.0015 (n = 69,663) |

No consistent pattern across datasets; some PTB-XL cells have fewer than 10 positives. Sex is the sex recorded in each dataset; PTB-XL ages above 89 years (coded 300) were counted as ≥ 65.

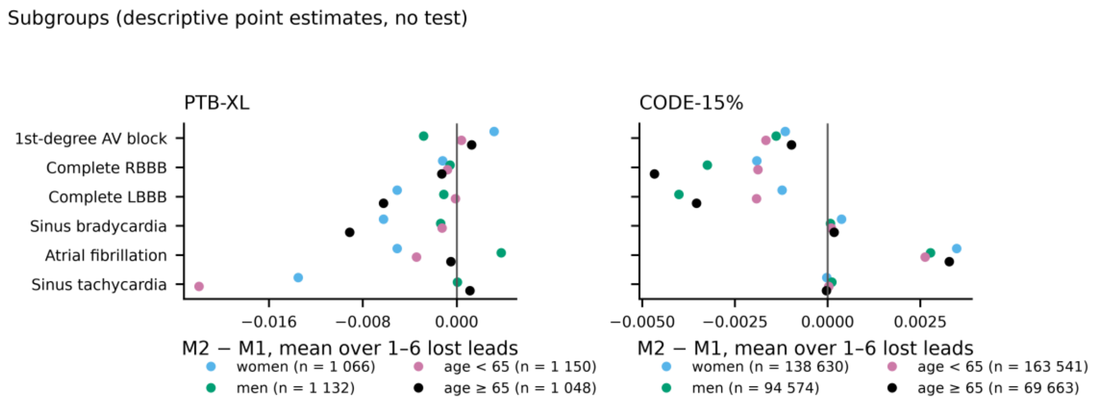


*Subgroups.*

### S12. Leakage control

With permuted training labels (M0, seed 0, 8 epochs), fold-9 macro-AUROC stayed at chance level (mean 0.534, range 0.478 to 0.591), against 0.979 with true labels.
