## Supplementary material for "Teaching an ECG model where to look does not make it more robust to missing leads: a pre-registered comparison of guideline-based attention guidance with random lead masking": TRIPOD-AI_checklist

Collins GS, et al. BMJ 2024;385:e078378. Item labels are abbreviated; “Main” = main manuscript (English version), “Suppl” = Supplementary Material. D = development, E = evaluation.

| Item | Topic (abbreviated) | Where reported |
|---|---|---|
| 1 | Title | Title page (identifies development and evaluation of deep-learning models, target population, outcomes) |
| 2 | Abstract | Abstract |
| 3a | Background, rationale | Introduction ¶1–3 |
| 3b | Target population, intended use, users | Introduction ¶1; Discussion 4.4 (clinicians, developers, evaluators) |
| 3c | Known inequalities between groups | Discussion 4.5; Suppl S11 (sex and age, descriptive) |
| 4 | Objectives | Introduction ¶4 (H1–H4) |
| 5a | Data sources | Methods 2.2 |
| 5b | Dates of data | PTB-XL recorded 1989–1996 (Germany); CODE-15% 2010–2016 (Brazil) — Methods 2.2 |
| 6a | Setting | Methods 2.2 (hospital and telehealth ECGs) |
| 6b | Eligibility | Methods 2.2 (10-s tracings; first exam per patient in CODE-15%) |
| 6c | Treatments | Not applicable (ECG classification) |
| 7 | Pre-processing, quality checks | Methods 2.3 |
| 8a | Outcome definition | Methods 2.2 (label rules) |
| 8b | Outcome assessors | Methods 2.2 (cardiologist statements in PTB-XL; partly automatic labels in CODE-15%); Limitations |
| 8c | Blinding of outcome assessment | Labels created before and independently of this study |

| Item | Topic (abbreviated) | Where reported |
|---|---|---|
| 9a | Choice of predictors | Methods 2.4 (raw 12-lead signal) |
| 9b | Predictor definitions | Methods 2.3–2.4 |
| 9c | Predictor assessors | Not applicable (signals) |
| 10 | Sample size | Methods 2.2; Table 1 (fixed public datasets; precision shown by CI width) |
| 11 | Missing data | Methods 2.2 (7-s tracings excluded); lead loss is the object of the study |
| 12a | Data use and partition | Methods 2.2 (official patient-level folds) |
| 12b | Handling of predictors | Methods 2.3 |
| 12c | Model type, building, tuning, internal validation | Methods 2.4–2.5; Suppl S2 (λ on fold 9) |
| 12d | Heterogeneity across clusters | Patient-level bootstrap; external dataset — Methods 2.7 |
| 12e | Performance measures, model comparison | Methods 2.7 |
| 12f | Model updating | None; recalibration recommended (Discussion 4.4) |
| 12g | Prediction calculation at evaluation | Methods 2.6–2.7 (five-seed ensemble) |
| 13 | Class imbalance | No rebalancing (binary cross-entropy); AUROC and calibration reported |
| 14 | Fairness | Suppl S11 (descriptive sex and age subgroups) |
| 15 | Model output, thresholds | Probabilities; no threshold (AUROC, calibration) |
| 16 | Development vs evaluation differences | Methods 2.2; Results 3.7; Limitations |
| 17 | Ethics | Declarations |
| 18a | Funding | Declarations |

| Item | Topic (abbreviated) | Where reported |
|---|---|---|
| 18b | Conflicts of interest | Declarations |
| 18c | Protocol | OSF doi:10.17605/OSF.IO/M9UR2 |
| 18d | Registration | OSF doi:10.17605/OSF.IO/M9UR2 |
| 18e | Data sharing | Declarations (public datasets) |
| 18f | Code sharing | Declarations (MIT licence, released with the article) |
| 19 | Patient and public involvement | None (public de-identified data) |
| 20a | Participant flow | Methods 2.2; Table 1 |
| 20b | Participant characteristics | Table 1 |
| 20c | Development vs evaluation characteristics | Table 1 |
| 21 | Numbers and events per analysis | Table 1; Suppl S7 |
| 22 | Full model specification | Methods 2.4–2.5; code released |
| 23a | Performance with CI, subgroups | Results 3.2–3.7; Table 3; Suppl S6–S11 |
| 23b | Heterogeneity of performance | Results 3.7; Suppl S7, S11 |
| 24 | Model updating results | Not applicable |
| 25 | Interpretation | Discussion 4.1–4.3 |
| 26 | Limitations | Discussion 4.5 |
| 27a | Poor-quality or missing input data | Object of the study (Methods 2.6) |
| 27b | User interaction, expertise | Discussion 4.4 |
| 27c | Future research | Discussion 4.6 |
